# Impact of climatic, demographic and disease control factors on the transmission dynamics of COVID-19 in large cities worldwide

**DOI:** 10.1101/2020.07.17.20155226

**Authors:** Soeren Metelmann, Karan Pattni, Liam Brierley, Lisa Cavalerie, Cyril Caminade, Marcus S C Blagrove, Joanne Turner, Kieran J Sharkey, Matthew Baylis

## Abstract

We are now over seven months into a pandemic of COVID-19 caused by the SARS-CoV-2 virus and global incidence continues to rise. In some regions such as the temperate northern hemisphere there are fears of “second waves” of infections over the coming months, while in other, vulnerable regions such as Africa and South America, concerns remain that cases may still rise, further impacting local economies and livelihoods. Despite substantial research efforts to date, it remains unresolved as to whether COVID-19 transmission has the same sensitivity to climate and seasonality observed for other common respiratory viruses such as seasonal influenza. Here we investigate any empirical evidence of seasonality using a robust estimation framework. For 304 large cities across the world, we estimated the basic reproduction number (R_0_) using logistic growth curves fitted to cumulative case data. We then assessed evidence for association with climatic variables through mixed-effects and ordinary least squares (OLS) regression while adjusting for city-level variation in demographic and disease control factors. We find evidence of association between temperature and R_0_ during the early phase of the epidemic in China only. During subsequent pandemic spread outside China, we instead find evidence of seasonal change in R_0_, with greater R_0_ within cities experiencing shorter daylight hours (direct effect coefficient = −0.247, p = 0.006), after separating out effects of calendar day. The effect of daylight hours may be driven by levels of UV radiation, which is known to have detrimental effects on coronaviruses, including SARS-CoV-2. In the global analysis excluding China, climatic variables had weaker explanatory power compared to demographic or disease control factors. Overall, we find a weak but detectable signal of climate variables on the transmission of COVID-19. As seasonal changes occur later in 2020, it is feasible that the transmission dynamics of COVID-19 may shift in a detectable manner. However, rates of transmission and health burden of the pandemic in the coming months will be ultimately determined by population factors and disease control policies.

## Introduction

An unusual pneumonia outbreak occurred in Wuhan, Hubei province, China in December 2019. The aetiological agent was found to be a novel coronavirus, and was named Severe acute respiratory syndrome coronavirus 2 (‘SARS-CoV-2’) (Gorbalenya et al., 2020). The associated disease, COVID-19, is believed to have originally spilled over from a wild animal to a human host, with the first cluster of cases linked epidemiologically to a seafood and wet animal wholesale market in Wuhan (Zhu et al., 2020). SARS-CoV-2 belongs to the *Betacoronavirus* genus, with high sequence homology to a wild bat coronavirus, pointing to bats as a likely natural reservoir (Guo et al., 2020). Since its initial discovery, COVID-19 has spread globally, with cases reported so far from 216 countries and territories. As of early July 2020, the COVID-19 pandemic remains a significant public health emergency, with global confirmed estimates exceeding ∼13.5 million cases and ∼584,000 deaths and global daily incidence is still increasing (WHO, 2020a).

After an incubation period that can range between two days and two weeks (5-7 days median), patients with COVID-19 typically present with fever, myalgia, cough, sore throat and difficulty breathing. However, asymptomatic infections are believed to occur, with a role in transmission that is still subject to debate (He et al., 2020; Moghadas et al., 2020; Treibel et al., 2020). SARS-CoV-2 is primarily transmitted through respiratory droplet but also by physical contact between hosts and to a lesser extent via fomite, similar to the common cold and influenza viruses (Wu et al., 2020).

Although epidemic transmission has been observed in virtually every climate type, respiratory infections such as human coronaviruses are often sensitive to effects of climate and weather. For example, the endemic human coronaviruses (*Alphacoronavirus:* Human coronaviruses 229E, NL63; *Betacoronavirus:* Human coronaviruses OC43, HKU1) follow established seasonal respiratory transmission in winter (Moriyama et al., 2020; Nickbakhsh et al., 2020). Previous epidemic coronaviruses, SARS-related coronavirus and Middle East respiratory syndrome (MERS)-related coronavirus have also exhibited associations between transmission and colder temperatures (Gardner et al., 2019; Lin et al., 2006). However, whether COVID-19 may be sensitive to climate remains a significant knowledge gap (Neher et al., 2020; Nickbakhsh et al., 2020). Governments and health authorities are increasingly concerned about observed rises in COVID-19 cases in southern hemisphere countries (e.g. Brazil, South Africa, Australia) since entering austral winter; and the possibility of resurgence (“second waves”) when northern hemisphere countries re-enter boreal winter at the end of 2020. There are also questions regarding whether seasonally-driven patterns of infection might continue longer-term, should COVID-19 persist endemically (Kissler et al., 2020; Neher et al., 2020). Assessing evidence for seasonality and climatic effects is therefore essential to help predict (and prepare for) potential short- and long-term climate-driven disease dynamics.

Early attempts to capture relationships between climate and case numbers or transmission rate have produced disparate and inconclusive results. Analyses have conflictingly reported positive (Jiang et al., 2020; Luo et al., 2020; Oliveiros et al., 2020), negative (Jüni et al., 2020; J. Wang et al., 2020; Ward et al., 2020), or nonlinear (Chen et al., 2020; Ficetola and Rubolini, 2020; Qi et al., 2020) effects of humidity. Reported effects of temperature appear more consistent, suggesting a consensus negative relationship (Hossain, 2020; Jiang et al., 2020; Luo et al., 2020; Oliveiros et al., 2020; Qi et al., 2020; Thangriyal et al., 2020; J. Wang et al., 2020), or curvilinear peaks of cases or transmission rate between ∼0°C and ∼10°C (Bannister-Tyrrell et al., 2020; Chen et al., 2020; Ficetola and Rubolini, 2020; Notari, 2020; Wan et al., 2020; M. Wang et al., 2020) although one such study suggested a positive temperature dependency up to a plateau of 3°C (Xie and Zhu, 2020). However, these initial modelling efforts often achieved relatively poor model fits to data, either because of study conducted at country or administrative division-level, ignoring heterogeneity in local climatic conditions and epidemic spread, or failure to account for additional confounding variables. A recent systematic review highlighted that 12 of 17 climatological analyses of COVID-19 did not include or discuss potential demographic or socioeconomic confounders (Mecenas et al., 2020). As COVID-19 transmission has been experienced globally and over only seven months worth of climatic variation, finding rigorous evidence for seasonal sensitivity brings challenges requiring a more carefully-considered methodology (Carlson et al., 2020).

Epidemics are typically characterised by exponential growth during their initial period and this growth rate can be used to calculate the basic reproduction number (R_0_), which is the number of secondary infections resulting from one infected individual in a fully susceptible population (Heffernan et al., 2005). Following the exponential growth, the epidemic slows down due to limitations on the availability of new susceptible individuals. In the present pandemic, this is largely caused by the imposition of control strategies. Here we adopt the logistic equation as a simple representation of this behaviour, capturing the initial exponential growth phase and continuing to describe the outbreak as the epidemic starts to slow. It is possible to parameterise models such as this by fitting against either incidence data (e.g., Pellis et al., 2020) or cumulative case data (Biswas et al., 2020; Biswas and Sen, 2020).

Here we show that for noisy data such as COVID-19 cases, fitting to cumulative data can provide more robust results than fitting to incidence data; we then use the fitted models to derive estimates of R_0_ for individual large cities (population > 0.5 million) across the world (n = 304, in 38 countries representing all permanently inhabited continents and a wide range of climatic conditions). Regression models are then used to explore potential effects of climatic factors upon R_0_ during epidemic growth while adjusting for national and city-level variation in demography, socioeconomics, and epidemic response.

This quantification of the relative influence of climate upon transmission is necessary to understand how the present pandemic may respond to approaching seasonal changes and likelihood of seasonal cycles or second waves of transmission.

## Methods

We defined a large city as having at least half a million people within population figures given by Cox (2019), considering greater metropolitan areas or agglomerations of neighbouring cities as one city. We then collected data from a number of sources that described cases for 321 of these large cities (Supplementary Material (SM), Table S1).

The exponential growth rate was estimated from the logistic population growth model fitted to cumulative case data. We fitted the logistic equation because it is numerically efficient in comparison to, for example, the SIR model, as it avoids the need for repeatedly solving a complex differential equation system and is just as effective in representing the early epidemic behaviour (Ma, 2020). We fitted cumulative data because we show here that this is a more robust method than using noisy incidence data (SM Section S2.1).

Our objective was to pick out the intrinsic exponential growth. We therefore focused on the initial data points prior to the control behaviour dominating the growth behaviour. Our algorithm (SM Section S2.2) automatically crops the data at the point of inflection where control starts to dominate over growth; an approach similar to that used by Hsieh and Chen (2009). The logistic model then accounted for any deviation from exponential growth prior to the point of inflection. The basic reproduction number, R_0_, was then calculated from the exponential growth rate following Wallinga and Lipsitch (2007) (SM Section S2.3). We excluded five cities for which model fits were considered unreliable due to fitting based on fewer than four days of incidence data, plus a further twelve cities for which the exponential growth phase started with more than 100 cases and thus did not capture R_0_ (SM Section S4.8).

We then assembled a set of covariates that may potentially explain variation in estimates of basic reproduction number, covering five broad categories: climatic, geographic, demographic, socioeconomic and epidemic response at city- or country-level resolution (SM Table S2, S2.4). Where no city-level data were available, country-level average data were substituted or imputed based on other covariates using a random forest-based procedure (Stekhoven and Bühlmann, 2012). Covariates that appeared strongly correlated (SM Figure S2) were discarded (preferentially retaining city-resolution covariates over country-resolution covariates) until the remaining set of input covariates did not demonstrate multicollinearity (i.e., all variance inflation factor values < 5).

To quantify associations between R_0_ and potential explanatory covariates, ordinary least squares (OLS) regressions were firstly constructed, taking full covariate sets and reducing to a minimal model by stepwise removal based on Akaike Information Criterion (AIC) score. Next, mixed-effects regression models were constructed by adding country-level random intercepts and model fit compared through likelihood ratio tests (LRT). In all regressions, cities were assigned weights proportional to the number of days of incidence data used to calculate exponential growth; the weights act as a proxy for confidence in the R_0_ estimate. To compare categories of covariates, relative contribution to the proportion of variance explained (R^2^) for each was calculated using proportional marginal variance decomposition (Grömping, 2007).

## Results

We applied the logistic model to cumulative data from 321 cities with available case data, and obtained reliable estimates of R_0_ for 304 cities with sufficient case data in their early epidemic (SM Sections S4.1 to S4.8). Resulting COVID-19 R_0_ values followed a slightly skewed distribution with median of 2.14 and interquartile range (IQR) of 0.69 (Table 1, Figure 1A), slightly lower than the previously reported median of 2.79 estimated from meta-analyses of twelve studies (ten in China) carried out in February 2020 (Liu et al., 2020). Observed values tended to be greater in cities within China than those within other parts of the world (Table 1, Figure 1).

**Table 1:** Descriptive statistics for values of R_0_ over exponential growth period for 304 world cities with at least half a million inhabitants, stratified by region. IQR = interquartile range.

| **Continent** | **Number of cities** | **Min** | **Median** | **Max** | **IQR** |
| --- | --- | --- | --- | --- | --- |
| China | 82 | 1.61 | 2.46 | 4.71 | 0.64 |
| South America | 23 | 1.48 | 2.41 | 3.89 | 1.10 |
| Australia | 1 | 2.05 | 2.05 | 2.05 | 0.00 |
| North America | 56 | 1.20 | 2.01 | 4.06 | 0.38 |
| Europe | 40 | 1.59 | 2.01 | 3.44 | 0.58 |
| Africa | 9 | 1.23 | 1.95 | 3.26 | 0.94 |
| Asia (exc. China) | 93 | 1.29 | 1.94 | 6.21 | 0.77 |
| *World* | *304* | *1.20* | *2.14* | *6.21* | *0.69* |

**Figure 1:**
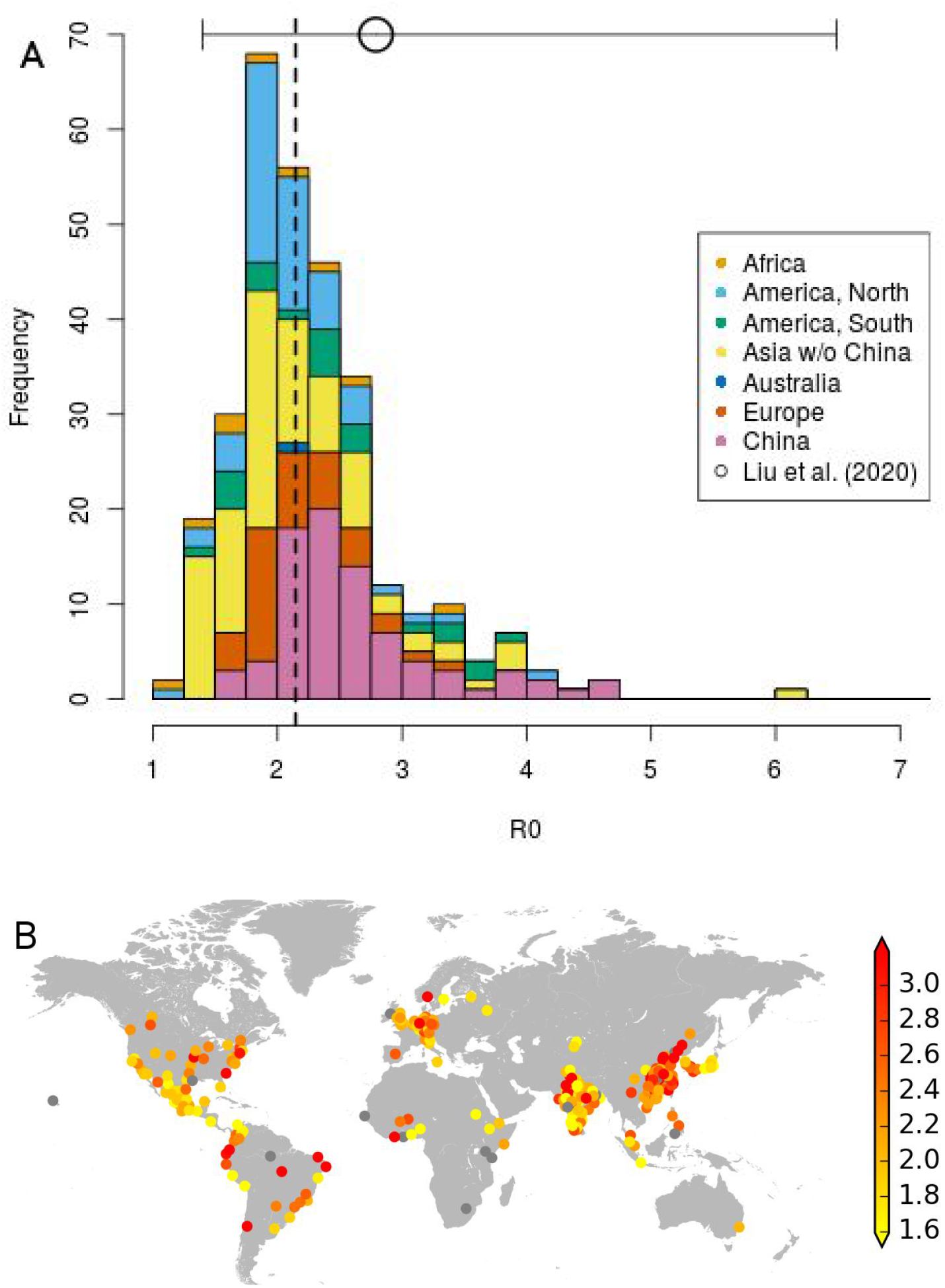
Calculated values of R_0_ over the exponential growth period for world cities with at least half a million inhabitants. A) Frequency histogram of R_0_ values calculated for 304 cities. Colour indicates continent, with China highlighted separately. Dashed line indicates median R_0_ value. The circle with horizontal bars indicates median, min and max of R_0_ values, as collected by Liu et al. (2020). B) Mapped locations of all 321 cities examined. Colour indicates R_0_ value, with grey dots indicating excluded cities (see SM S4.8).

A further three cities (Mexico City and Guadalajara, Mexico; Udaipur, India) were excluded during regression modelling based on excessive influence over model fits determined by Cook’s distance, Udaipur having the highest R_0_ estimate of 6.21. Initial models using data from the remaining 301 cities demonstrated an effect of daylight (and to some extent, stringency of government response) upon R_0_ (SM Figure S3A). As China was the centre of the initial phase of the pandemic when daylight hours in the northern hemisphere were shortest, and also represented over a quarter of cities within our data, regression analyses were constructed separately for a) global data excluding China (n = 219 cities) and b) China only (n = 81, further excluding Zhongshan based on Cook’s distance).

Considering global cities outside China, climate variables (including daylight) did not appear to explain variation in R_0_; evidence was instead observed for association with stringency of government response and GDP (SM Table S3). Introducing random intercepts significantly improved model fit (LRT statistic = 19.28, df = 1, p(LRT) = < 0.001), as estimated R_0_ values showed slightly greater clustering within-country than between-countries (intraclass correlation = 0.024). Following stepwise selection, the final fitted model retained stringency of government response as well as population size and density, but did not retain GDP after adjusting for this country-level clustering (Table 2). This model explained only 5.8% of variance in estimated R_0_ and did not fit particularly better/worse by continent (Figure 2), though consistently under-predicted for cities with elevated R_0_ values (> 3), including three cities in Brazil, seven in India, and three in the United States.

**Table 2:**
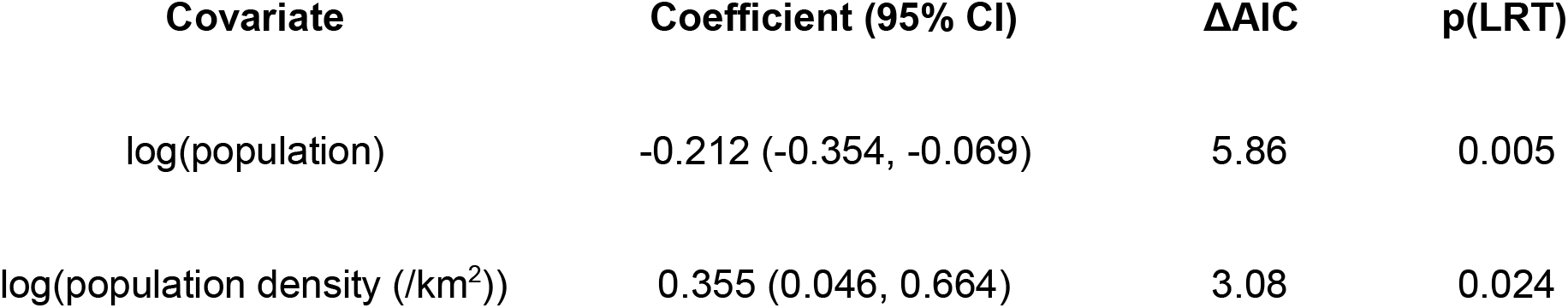
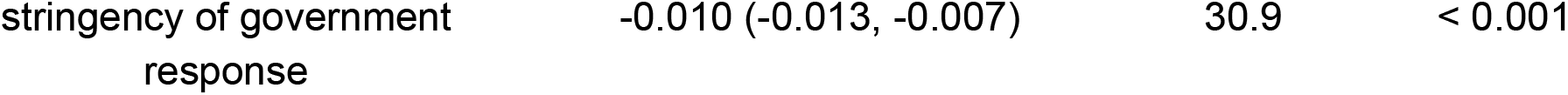
Outputs from selected mixed-effects regression model predicting R_0_ within global cities excluding China (n = 219) based on stepwise reduction from saturated model using AIC. CI = confidence interval, ΔAIC = change in Akaike Information Criterion when term excluded, LRT = Likelihood ratio test.

**Figure 2:**
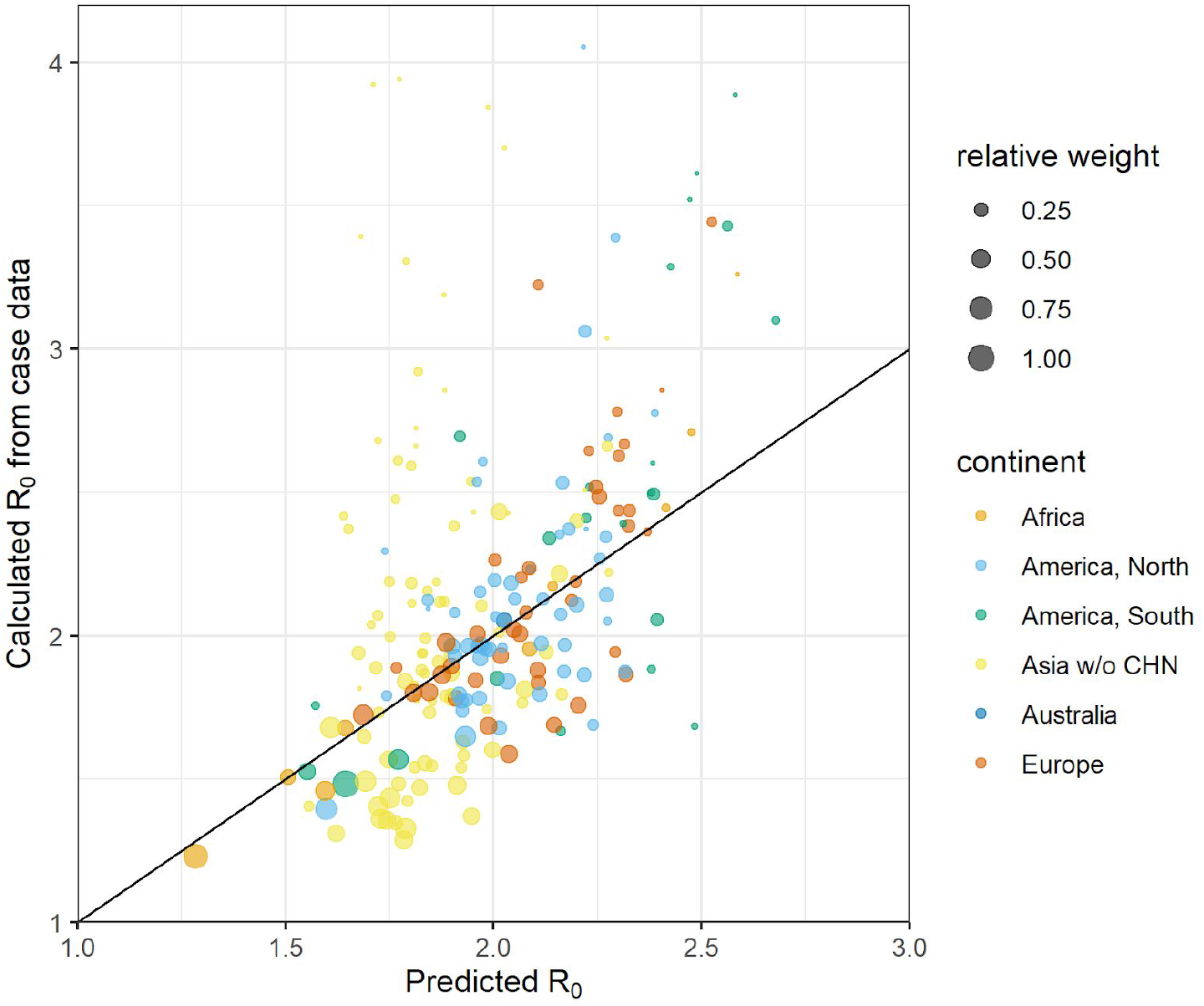
Plotted model performance for selected mixed-effects regression model predicting R_0_ in global cities excluding China (n = 219) based on demographic and epidemic response covariates. Size of points is proportional to weighting in model, determined as number of observed available days of incidence. Cities above diagonal have under-estimated R_0_, while those below have over-estimated R_0_.

Adjusted for other covariates, R_0_ was greater in cities with a smaller, but more dense population (Table 2 and Figure 3). Additionally, impact of control measures may be reflected in the observed association with mean stringency of government response two weeks prior; for a score increase of 10 in stringency index (measured from 0 – 100; see Hale et al. (2020)), we observed an average decrease of 0.10 in estimated R_0_ (Figure 3C).

**Figure 3:**
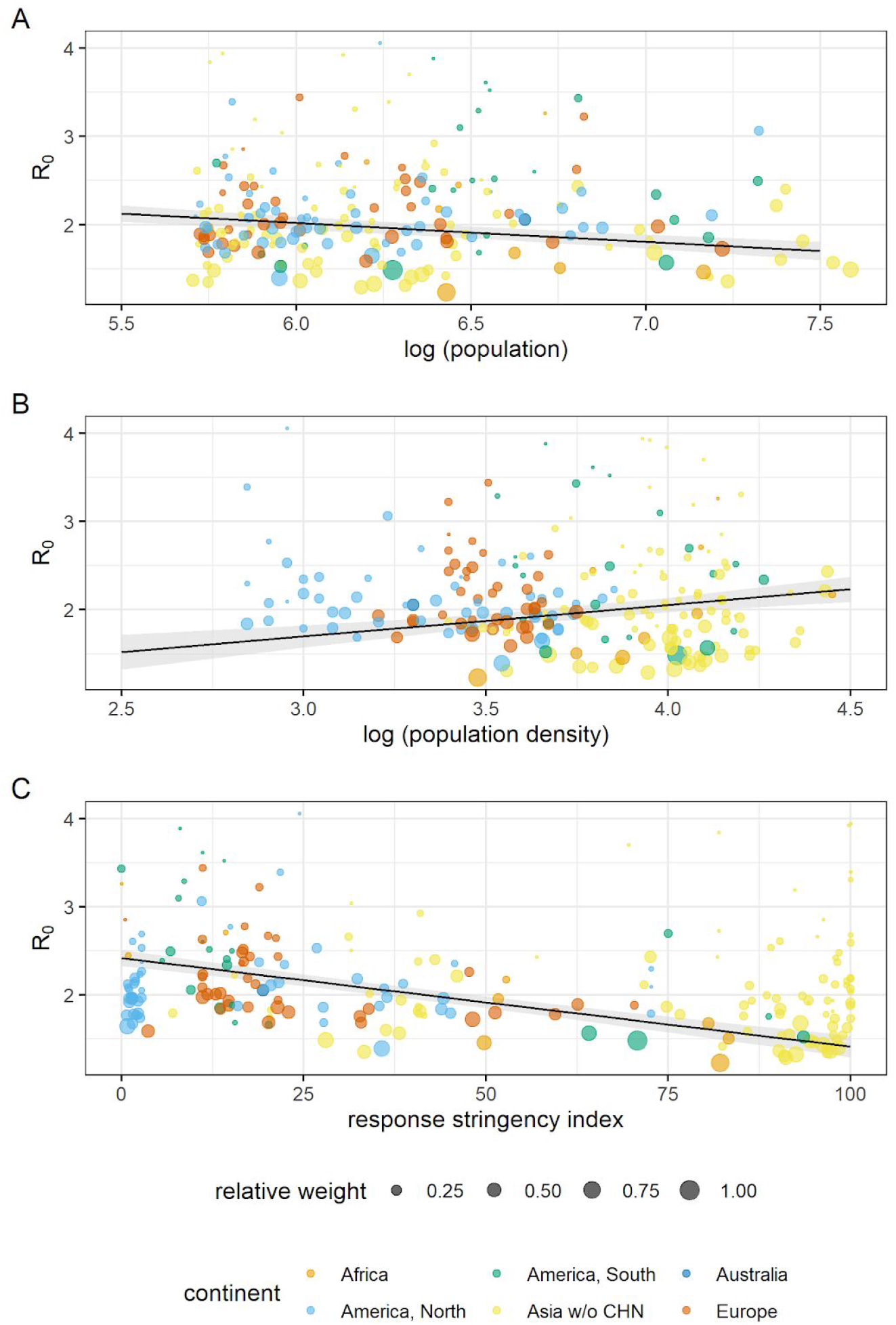
Plotted covariates from selected mixed-effects regression model predicting R_0_ within global cities excluding China (n = 219), showing effect of A) population size, B) population density and C) index measuring stringency of government response two weeks before epidemic growth period. Size of points is proportional to weighting in model, determined as number of observed days of incidence. Lines denote fits, calculated as estimated marginal means holding all other model variables constant. Shaded areas denote 95% confidence interval.

Apparent effects of daylight were no longer observable in the global data excluding cities in China. As daylight hours is a function of calendar day, these initial effects may have simply been driven by differences in R_0_ between early emergence and the later pandemic phase. Therefore, we investigated whether there was any evidence for seasonal effects independent of calendar day by conducting mediation analysis using dates of epidemic growth period (SM Section S3.3). We observed a significant, negative, direct effect of daylight hours (coefficient = −0.247, p = 0.006), and a significant, positive, indirect effect of daylight via calendar day (coefficient = 0.158, p < 0.001) (SM Table S4), offering support to both hypotheses of seasonality in COVID-19 spread and effects of outbreak phase in time. Notably, the direct effect of daylight in mediation analysis was slightly stronger than in the initial model fitted to 301 global cities (SM Figure S3A); opposite signs between direct and indirect effects implied some antagonism between daylight and calendar day effects.

When considering cities in China only, regression models were constructed excluding any covariates with country-level data (i.e., constant values for cities), multicollinearity or consisting of imputed data only, leading to exclusion of daylight hours, elder ratio, GDP per capita, air pollution and change in retail and recreation (SM Table S5).

In the China-specific model, R_0_ was associated with only two covariates representing climate and epidemic response (Table 3) which collectively explained 8.7% of variation in R_0_. We observed evidence that warmer climates within China experienced lower rates of transmission; within the final fitted model, estimated R_0_ decreased by an average of 0.25 for every 10°C increase (Figure 4A). We additionally observed only tentative evidence for an association with mean stringency of government response two weeks prior (Figure 4B), though in a positive direction, potentially indicating difficulty of controlling the early epidemic.

**Table 3:** Outputs from selected regression model predicting R_0_ within cities in China (n = 81) based on stepwise reduction from saturated model using AIC. CI = confidence interval, ΔAIC = change in Akaike Information Criterion when term excluded, LRT = Likelihood ratio test.

| Covariate | Coefficient (95% CI) | ΔAIC | p(LRT) |
| --- | --- | --- | --- |
| temperature (°C) | -0.025 (-0.048, -0.002) | 2.54 | 0.033 |
| stringency of government response | 0.012 (-0.001, 0.025) | 1.21 | 0.073 |

**Figure 4:**
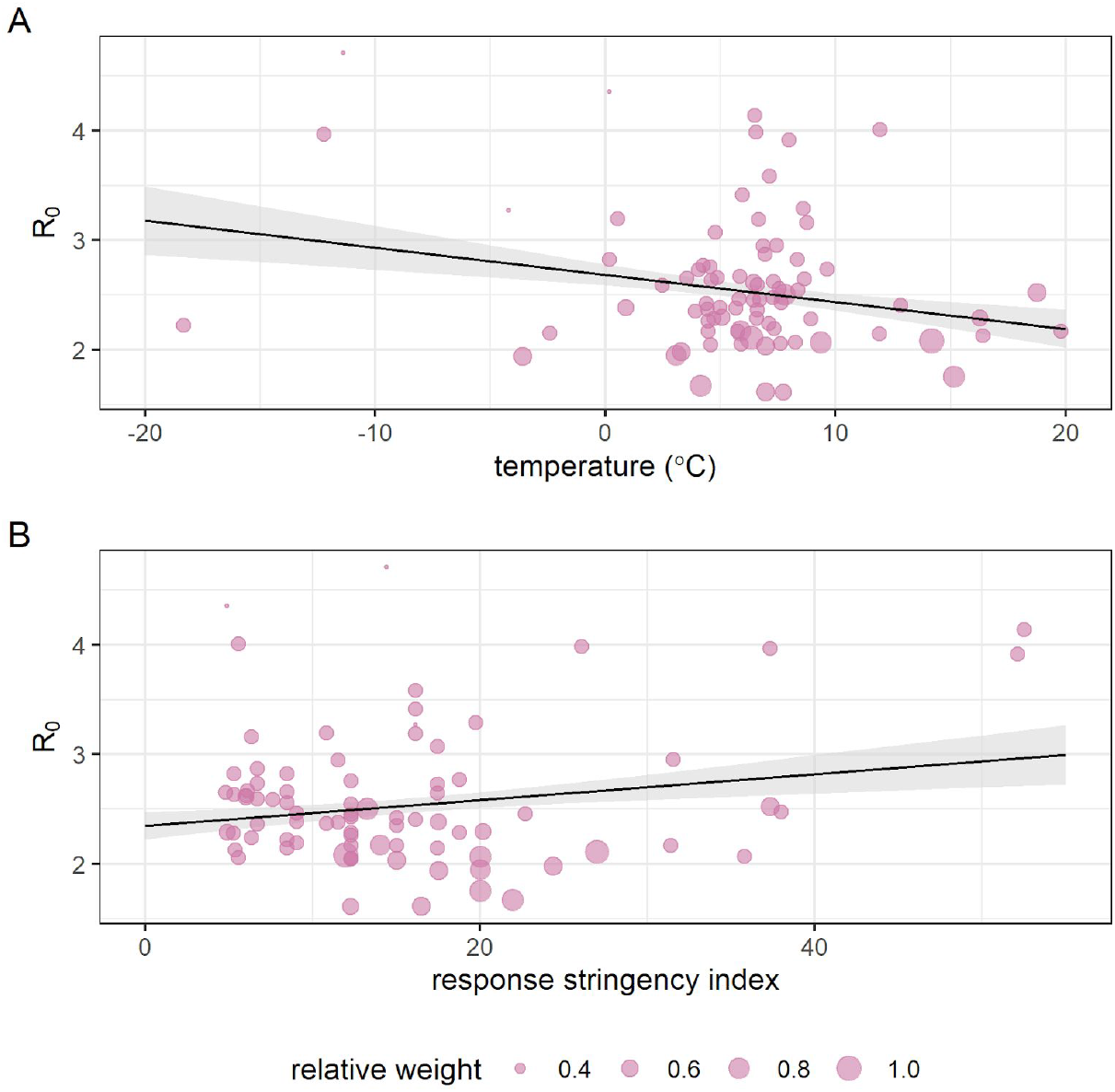
Plotted covariates from selected regression model predicting R_0_ within cities in China (n = 81), showing effect of A) mean daily temperature, and B) index measuring stringency of government response two weeks before epidemic growth period. Lines denote fits, calculated as estimated marginal means holding all other model variables constant. Shaded areas denote 95% confidence interval.

Although overall explanatory power was limited in all cases, a comparison of saturated models (SM Tables S3 and S5) confirmed climate variables had greater relative explanatory power of R_0_ estimates in China than the rest of the world (Figure 5). Contrastingly, demographic and epidemic response covariates explained the greatest proportion of variance in R_0_ estimates for our global analysis but were less relatively informative within analysis of Chinese cities only.

**Figure 5:**
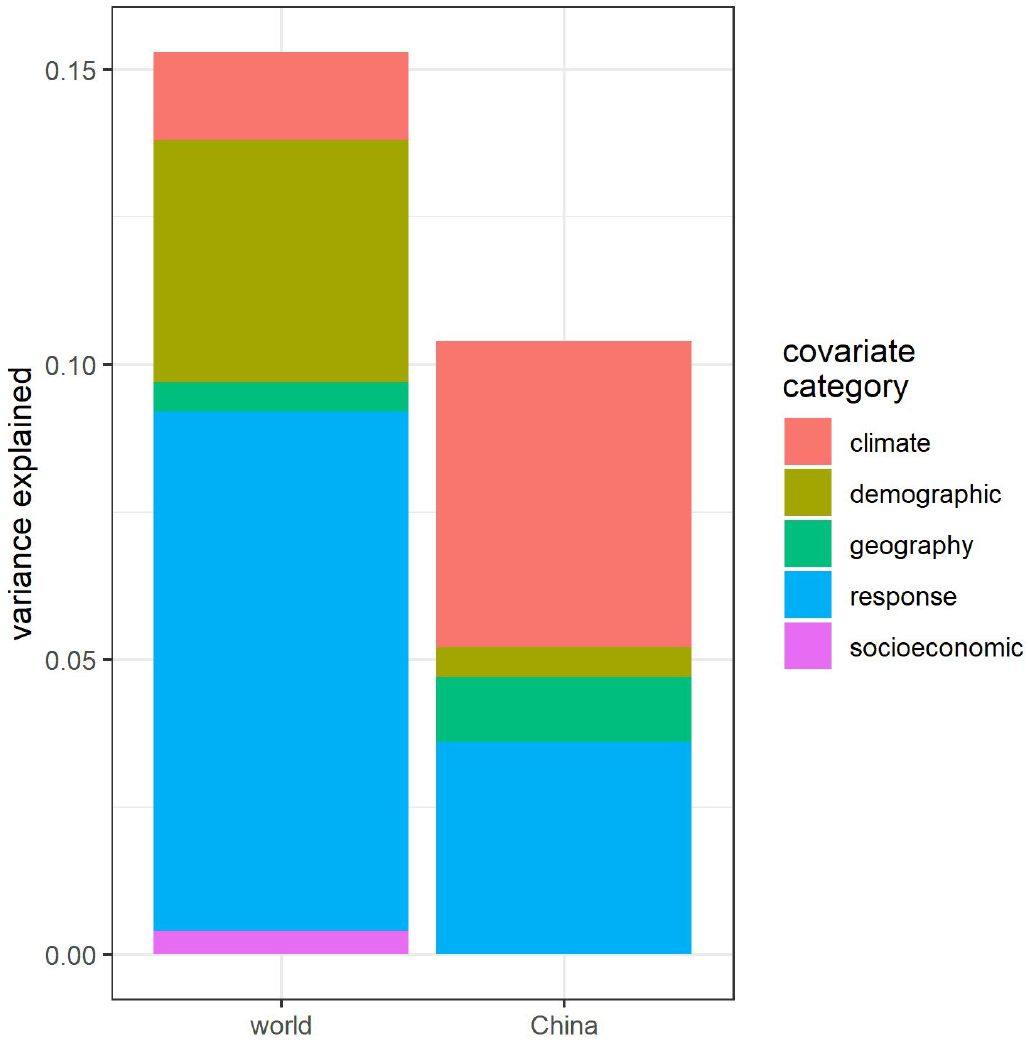
Relative importance of each covariate category in saturated regression models predicting R_0_ within global cities excluding China, and cities within China only. Y axis denotes collective proportion of variance explained, i.e., contribution to model R^2^. See SM Table S2 for explanation of covariate categories.

## Discussion

Although COVID-19 achieved pandemic spread within a short period of time, the extent of transmission has greatly varied across large metropolitan areas worldwide (Figure 1). We observe the most intense exponential growth periods and highest estimated R_0_ values primarily within China but also several other Asian, and North and South American cities. We show that temperature may tentatively explain variation in R_0_ in the early epidemic in China (Table 3 and Figure 4), but did not explain such variation as the pandemic spread worldwide, when demographic and epidemic response factors were more influential (Table 2 and Figure 3). However, we observed support for climatic effects through effects of daylight after filtering out mediation from calendar day. Finally, we also report an estimable impact of governmental responses in association between R_0_ and strictness of disease control at a global scale.

To determine the underlying exponential growth rate of the epidemic, we fit the logistic equation to the total cumulative number of cases, rather than the more standard approach of fitting to daily incidence data. This leads to more robust results, confirmed by our comparisons in SM Section S2.1 where we also observe an upward bias in growth rate when using incidence data. Fitting to cumulative data is known to cause correlated errors (King et al., 2015; Ma et al., 2014), but for our purposes, using cumulative data gives a more reliable estimate of the growth rate (see also King et al., 2015). The R_0_ values are calculated using a mean serial interval of 4.7 days for SARS-CoV-2 (Nishiura et al., 2020). Our R_0_ estimates obtained for China (median = 2.46) are consistent with the meta-review study of Liu et al. (2020) (median = 2.79, range = 1.40 - 7.23) and other recent review studies (Alimohamadi et al., 2020; Park et al., 2020). Liu et al.(2020) highlighted that SEIR type compartmental models produce higher estimates of R_0_ (for example, Tang et al. (2020), range = 5.7 - 7.2).

One of the most hotly debated questions is whether transmission of COVID-19 is affected by climate, and therefore, whether we can anticipate R_0_ to be affected by forthcoming seasonal changes (Neher et al., 2020; Nickbakhsh et al., 2020). Considering data exclusively from China, although overall explanatory power was low, we observe an effect of temperature suggestive of higher R_0_ in colder cities (Table 3 and Figure 4A). Similarly, a systematic review supported significant trends in COVID-19 incidence or transmission with temperature (Mecenas et al., 2020) with increases in colder conditions reported in 16 of 17 studies (e.g., Bannister-Tyrrell et al., 2020; Luo et al., 2020; Oliveiros et al., 2020; J. Wang et al., 2020). Comparing to other analyses using linear regression methods to explain R_0_, our estimated effect size of temperature (β = −0.025) is smaller than reported in a pooled study of East Asia (Luo et al. (2020): −1.050) but similar to those of a study that also stratified by country and adjusted for sociodemographic factors (J. Wang et al. (2020): China: −0.023; USA: −0.020). Decreases in transmission in more humid conditions were reported by 13 of 14 studies (e.g., Chen et al., 2020; J. Wang et al., 2020) but we find no evidence of an association between humidity and R_0_. Overall quality of evidence for these climatic associations in the systematic review was noted to be low (Mecenas et al., 2020).

Several of these studies pooled global data. We find that such pooling of global data may be inappropriate if data represent different pandemic phases in calendar time. Exponential growth in cases mostly occurred in January for cities within China and in March - May elsewhere, with a marked contrast in research knowledge and policymaker awareness between these periods. Previously reported effects using global pooled data may therefore have additionally attributed differences in epidemic preparedness to climatic effects upon R_0_, leading to overestimation of the importance of climate.

When considering data from the pandemic phase outside of China, we find evidence of greater values for R_0_ within cities experiencing shorter daylight hours, whilst adjusting for disease control measures and mediation from time of year (SM Table S4). COVID-19 transmission is, therefore, sensitive to a climatic variable, and one which changes with season, particularly at further distance from the equator. However, the effect is small and may be swamped by greater sensitivity of COVID-19 to epidemic responses and demography (Figure 3 and 5). This finding offers empirical support to a recent scenario analysis using mathematical models (Baker et al., 2020), which predicted the influence of climate upon COVID-19 transmission to be minimal compared to supply of susceptibles - a function of epidemiological dynamics and efficacy of control measures. This is also coherent with more recent statistical modelling efforts finding climatic variables to be of less importance compared to, for example, the number of public health interventions (Jüni et al., 2020), and airport connectivity (Coelho et al., 2020).

If not temperature or humidity-linked, then daylight hours might feasibly impact the epidemiology of COVID-19 through ultraviolet (UV) radiation. UV light is considered one of the possible drivers behind the seasonality of respiratory viruses such as influenza and common cold viruses (Li et al., 2019), among human behavioural factors (i.e. indoor crowding during colder days) and physiological factors (i.e. impacts of colder seasonal weather on human immunity (Eccles, 2002)). UV light has been hypothesised to have effects on community transmission through viral inactivation (Sagripanti and Lytle, 2020). Higher UV radiation has been significantly associated with lower average COVID-19 growth rate in grid-scale analysis (Byass, 2020) and between countries or states (Carleton et al., 2020; Merow and Urban, 2020), though only later in the year in the latter analysis, broadly mirroring the daylight effect in our models excluding early epidemic data from China.

Consistent with our findings, early experimental evidence for SARS-CoV-2 appears to show minimal effect of temperature on virus longevity (although a slight peak in longevity at 30°C is reported; Kratzel et al., 2020). This contrasts with convincing experimental evidence linking temperature and humidity to the viability of SARS-CoV-1. Chan et al. (2011), for example, showed stable viability of dried virus for over five days at 22-25°C and RH of 40-50%; whilst at 38°C, and RH of >95%, a >2 log reduction in viral titre was observed in just 7 hours.

UV light has complex experimental effects on coronaviruses. The major component of UV in sunlight, UVA, has minimal effectiveness upon many viruses (Kowalski, 2009; Pozo-Antonio and Sanmartín, 2018), including SARS-CoV-1 (Darnell et al., 2004). UVC is commonly used as a virucidal and effectively inactivates SARS-CoV-1 (Darnell et al., 2004), but is unlikely to have impacts in non-laboratory settings as only minimal UVC passes the ozone layer (Kowalski, 2009). However, UVB, which is present in small amounts in natural sunlight, rapidly inactivates SARS-CoV-2 on surfaces (Ratnesar-Shumate et al., 2020). Simulated sunlight representative of midday on the summer solstice at 40°N latitude resulted in a 90% reduction of viable virus every 6.8 minutes in simulated saliva; whilst sunlight representative of winter solstice at the same latitude required 14.3 minutes for a 90% reduction. Furthermore, no significant decay was observed in darkness over 60 minutes (Ratnesar-Shumate et al., 2020), consistent with other data on both SARS-CoV-2 and SARS-CoV-1 (Rabenau et al., 2005; van Doremalen et al., 2020). These laboratory experiments are consistent with our study’s findings of a negative association between R_0_ and daylight hours, which may be explained by the virucidal effect of UV radiation.

The presented analytical framework is subject to some methodological assumptions and caveats. Firstly, our study relies on six months of case data with variable data quality between sources. For example, data describing case counts in China between different sources have demonstrated some inconsistencies (Python et al., 2020), potentially as a result of differences in case definitions or magnitude and strategy of testing. Similarly, epidemic trajectory is still currently unclear for several cities, particularly those in Africa where cases may not have yet reached peak epidemic growth (WHO, 2020b).

A more detailed analysis of climate variability would require data at interannual to decadal time scales, though only one previous betacoronavirus pandemic has occurred to date. For influenza, Shaman and Lipsitch (2013) have highlighted that each major pandemic (1918, 1957, 1968, and 2009) was preceded by La Niña conditions e.g. colder sea surface temperatures than average in the equatorial Pacific. This year, winter temperature conditions were close to neutral in the Pacific, but a mild La Niña signal seems to be developing during boreal spring-summer 2020 (NOAA Climate Prediction Center, 2020) and further data will be necessary for formal influenza comparisons.

Our analysis covers large cities on all inhabited continents. However, data was unavailable for cities in the arid Middle East and in colder parts of the world, e.g. northern Russia, therefore our models represent a restricted part of the full climatic range of human inhabited regions. Our data are also substantially biased towards countries exhibiting northern hemisphere patterns of seasonality (n = 143 cities above 30° latitude compared to n = 4 cities below −30°). Finally, the models we present explain only a low proportion of variance of our estimated R_0_ values, a somewhat expected result for ‘ecological’ global-resolution correlative analyses. The rate of spread of COVID-19 is likely to be subject to many other factors not possible to include here, including detailed heterogeneity in population structure or movements and varying individual susceptibility to infection.

Our results support the prudent approach of WHO and CDC public-facing guidelines to not raise expectations that the COVID-19 outbreak will stop as a result of warmer weather. On the contrary, there are increasing concerns about “second waves” of epidemic spread during winter (Academy of Medical Sciences, 2020), i.e. as daylight hours decrease. Of particular importance will be compliance with lockdown restrictions and increased testing to ensure effective disease control.

Simulations show that seasonal sensitivity of COVID-19 could plausibly lead to recurrent outbreaks each winter (Kissler et al., 2020). Our observed evidence for daylight effects and other experimental evidence regarding virucidal capabilities of UV light would support the possibility of such a scenario. However, we find weaker evidence for a link between temperature and R_0_, concordant with a lack of increased molecular viral longevity under cooler experimental conditions observed elsewhere, reducing support for the likelihood of long-term seasonal cycling. If COVID-19 becomes endemic, movement of infected individuals from the northern or southern hemisphere to other parts of the world in their respective (boreal or austral) winters may be considered a speculative driver of future epidemic recurrence to be monitored carefully.

Although we focus primarily on northern hemisphere patterns, equal concern has been raised regarding the southern hemisphere, as epidemic spread is resurging in Australia and continues to be very active in Madagascar and South Africa. More widely, Africa hosts 16% of the global population, yet to date accounts for just 2.9% of confirmed COVID-19 cases and 1.2% of reported deaths (WHO, 2020b). Higher temperature has been put forward as an explanatory hypothesis (Njenga et al., 2020), which our results do not support. Although the annual range in temperature is small, equatorial Africa may yet demonstrate more complex seasonal patterns of COVID-19, from no seasonality to several epidemics per year, as shown for influenza dynamics in Kenya (Emukule et al., 2016; WHO, 2017).

Ultimately, our work implies that seasonal change in climate may have detectable effects upon forthcoming patterns of COVID-19 transmission, but that demographic and epidemic response factors will be much more influential. As a result, it is most prudent for policymakers to focus attention on disease control measures at this stage (O’Reilly et al., 2020).

## Conclusion

The assumption that COVID-19 will follow seasonal patterns as for endemic human coronaviruses and other respiratory illnesses is not unfounded, but requires careful empirical consideration. By taking a standardised dataset of large cities, we estimate R_0_ using a robust methodology and find seasonality to be supported in temperature effects, but only restricted to China. Instead, we find more compelling evidence that seasonal change in daylight hours is negatively associated with R_0_, following pandemic spread beyond China. Although we focus here on identifying and quantifying any seasonal component to COVID-19 transmission, we reiterate the importance of demography and epidemic responses in COVID-19 epidemiology. We cannot control how the seasons change, but we can control our immediate disease prevention measures and policies and we urge governments to remain vigilant and evidence-informed in all climates, particularly so during upcoming winter seasons with lower daylight conditions.

## Data Availability

Detailed citations and links for all used data are given in the SM.

## Acknowledgements

SM and KP acknowledge funding support from the NIHR Health Protection Research Unit in Emerging and Zoonotic Infections, and the Centre of Excellence in Infectious Diseases Research, and the Alder Hey Charity, awarded through the Liverpool COVID-19 Partnership Strategic Research Fund. The NIHR Health Protection Research Unit in Emerging and Zoonotic Infections is a partnership between the University of Liverpool, Public Health England (PHE) and Liverpool School of Tropical Medicine (LSTM). The views expressed are those of the authors and not necessarily those of the NHS, the NIHR, the Department of Health or Public Health England.

